# Modeling the potential impact of indirect transmission on COVID-19 epidemic

**DOI:** 10.1101/2021.01.28.20181040

**Authors:** Jummy David, Sarafa A. Iyaniwura, Pei Yuan, Yi Tan, Jude Kong, Huaiping Zhu

## Abstract

The spread of SARS-CoV-2 through direct transmission (person-to-person) has been the focus of most studies on the dynamics of COVID-19. The efficacy of social distancing and mask usage at reducing the risk of direct transmission of COVID-19 has been studied by many researchers. Little or no attention is given to indirect transmission of the virus through shared items, commonly touch surfaces and door handles. The impact of the persistence of SARS-CoV-2 on hard surfaces and in the environment, on the dynamics of COVID-19 remain largely unknown. Also, the current increase in the number of cases despite the strict non-pharmaceutical interventions suggests a need to study the indirect transmission of COVID-19 while incorporating testing of infected individuals as a preventive measure. Assessing the impact of indirect transmission of the virus may improve our understanding of the overall dynamics of COVID-19. We developed a novel deterministic susceptible-exposed-infected-removed-virus-death compartmental model to study the impact of indirect transmission pathway on the spread of COVID-19, the sources of infection, and prevention/control. We fitted the model to the cumulative number of confirmed cases at episode date in Toronto, Canada using a Markov Chain Monte Carlo optimization algorithm. We studied the effect of indirect transmission on the epidemic peak, peak time, epidemic final size and the effective reproduction number, based on different initial conditions and at different stages. Our findings revealed an increase in cases with indirect transmission. Our work highlights the importance of implementing additional preventive and control measures involving cleaning of surfaces, fumigation, and disinfection to lower the spread of COVID-19, especially in public areas like the grocery stores, malls and so on. We conclude that indirect transmission of SARS-CoV-2 has a significant effect on the dynamics of COVID-19, and there is need to consider this transmission route for effective mitigation, prevention and control of COVID-19 epidemic.

## Introduction

COVID-19 is a disease caused by the virus called severe acute respiratory syndrome coronavirus 2 (SARS-CoV-2), and was first identified in December 2019, which has since resulted into a pandemic. Despite the widely use of non-pharmaceutical interventions (NPIs), COVID-19 in Ontario, Canada and in many part of the world has been on the rise. Although several reports have shown that most people with mild COVID-19 illness will recover on their own [1, 2]. The spread of SARS-CoV-2 through direct transmission has been discussed in several reports and articles. The virus shed by infected individuals could also be contracted through indirect transmission pathway by touching contaminated surfaces or objects, and then touching of eyes, nose and or mouth [3–7]. The indirect transmission of SARS-CoV-2 was first reported in Wuhan, the capital of Hubei, China [8]. In addition, there have been reports that the airborne transmission of the virus via aerosols could also be a significant transmission pathway when in an enclosed system [9]. Recent measurements identified SARS-Cov-2 RNA on aerosols in Wuhan’s hospitals [10, 11], in a London hospital [12] and outdoor in northern Italy [13], unraveling the likelihood of indoor and outdoor airborne transmission of the virus [14, 15].

Many mathematical models considering direct transmission pathway have been developed to study the transmission, prevention and control of COVID-19 epidemic. These models have been used to reduce the risk of direct transmission using non-pharmaceutical interventions (NPIs). Models such as Yuan et al. [16] developed a SEAIR model to explore the efficacy of stay-at-home policy (SAHP), Xue et al. [17] developed a network transmission model to study COVID-19 epidemics in Wuhan (China), Toronto (Canada), and Italy, Wu et al. [18] estimated the trend of COVID-19 in Ontario and informed future actions for controlling the outbreak, Li et al. [19] developed a model to capture the transmission of the virus in Wuhan. None of these aforementioned models accounted for infections caused by indirect transmission and incorporating testing of individuals.

Considering the continuing increase in the number of cases of COVID-19, the debates on the transmission, and several episodes of the disease in the grocery stores, malls and public places, even with the implementation of social distancing and mask usage, we see that the current measures are not enough to eliminate the spread of COVID-19. Evidence suggests the need for further and in-depth studies on the transmission of COVID-19 through indirect route. For example, the indirect transmission of SARS-CoV-2 at the loblaw stores [20] and Sobeys [21] where staffs and customers are mandated to wear mask and keep to the physical distancing rule. Therefore, in order to reduce the risk of transmission and to implement additional preventive and control measures, there is need to look into how the disease is transmitted through indirect pathway. A few mathematical models such as the models in Yang et al. in [22] and Asamoah et al. in [23] incorporated multiple transmission routes to account for infections caused through both direct route (human-human) and indirect route (human-source-human). Yang et al. proposed a model using an endemic scenario with varying transmission rates without considering the impact of testing of infected individuals on the infection level. Similarly, Asamoah et al. developed a SEAIRV model to estimate the basic reproduction number with and without control using an optimal control theory. Neither Yang et al. nor Asamoah et al. incorporated an isolated and hospitalized classes to look into the role of shedding parameters on the peak of the infection, the reproduction number and the final epidemic size. In general, the most popular mathematical models discussed are the SEIR-type compartmental models.

In light of the continuous increase in the risk of COVID-19 transmission, its close association with indirect transmission, it is important to examine and understand the effect of shedding of viruses by infectious individuals on the epidemic of COVID-19 in Toronto while taking testing and isolation into consideration. Therefore, we developed a susceptible-exposed-infected-removed-virus-death (SEIRVD) deterministic model and incorporated infections transmitted through indirect pathways. The model differs from other existing model since we incorporated testing of both asymptomatic and symptomatic individuals. In addition, our model is the first to use multiple transmission routes to estimate the impact of indirect transmission on the peak, peak time, the effective reproduction number and the epidemic final size. We focus our analysis on identifying the role of indirect transmission on the disease burden and how lowering this transmission route through fumigation, disinfection could reduce the COVID-19 epidemic in Toronto and other similar settings. This work will provide important information, ideas and knowledge on how to improve current public health preventive and control measures and/or introduce new measures (new disinfection products) to reduce indirect transmission and achieve elimination targets in Toronto and other similar regions. Several scenarios with different parameters will be explored for appropriate generalizability of our results.

## Materials and methods

On January 19, 2021, Ontario province reported 1,913 new cases, with 46 new deaths, resulting into 242,277 cumulative confirmed cases with 5,479 total COVID-19-related deaths, and 209,183 recovered cases [24]. Similarly, on January 18, 2021, the city of Toronto reported 583 newly confirmed cases, with 5 new deaths, resulting into 75,273 cumulative confirmed cases with 2,211 total COVID-19-related deaths, and 67,219 recovered cases [25]. In addition to cases transmitted through close contacts, several evidences point to indirect transmission of COVID-19 in loblaw stores [20] and Sobeys [21], we present a SEIRVD model to study the effect of indirect transmission, specifically virus shedding while testing individuals, on COVID-19 prevention, control and elimination strategies to help improve public health policy in Toronto and other similar regions.

To capture the dynamics of COVID-19 epidemic in Toronto, we estimated some unknown parameters from model calibration based on the available data gotten from the Toronto Public Health (TPH) from March 1, 2020 to August 31, 2020. From the model and the estimated parameters, we analytically accessed the transmission risk using the computed effective reproduction number and final size relation. The reproduction number of the combined model is calculated using the next generational matrix approach [26, 27], and expressed as the sum of the effective reproduction numbers of the direct transmission and that of indirect transmission pathways. Similarly, the final size relation is estimated following the approaches in [28–30]. In addition, the developed model can be used to show the impact of viruses shed into the environment on the current and future epidemic trends, identify how indirect transmission increases the epidemic in Toronto, and as well suggest additional and new control measures.

### SEIRVD Model with indirect transmission

The mathematical model presented in this work is an SEIRVD-type compartmental model with no demographic effects. In this model *S* represents the susceptible population, *E* is the population of exposed individuals, *I*_*A*_ represents asymptomatic individuals (individuals with no symptoms), *I*_*S*_ is for symptomatic individuals (individuals with symptoms), *I*_*H*_ represents hospitalized individuals, *I*_*W*_ is the population of infected individuals self-isolating, *R* is the population of recovered individuals, and since there has not been many reported cases of reinfected individuals, the model ignores reinfection and assume that recovered individuals become immune to the virus. In the model, *V* is the amount of virus shed by infected individuals in the *I*_*A*_ and *I*_*S*_ compartments, and *D* is the population of deceased due to COVID-19. The model assumes that infected individuals in the compartments *I*_*H*_ (hospitalized) and *I*_*W*_ (isolated) neither transmit nor shed the virus since these individuals are confined in a particular place (home, hospital or isolation centre) with restricted movements and so do not spread the disease. Fig 1 shows the full transmission dynamics of the model with *λ* = (1 − *p*_1_) *S β*_*DA*_ *I*_*A*_ + (1 − *p*_1_) *S β*_*DS*_ *I*_*S*_ + (1 − *p*_2_) *S β*_*I*_ *V*.

**Fig 1.**
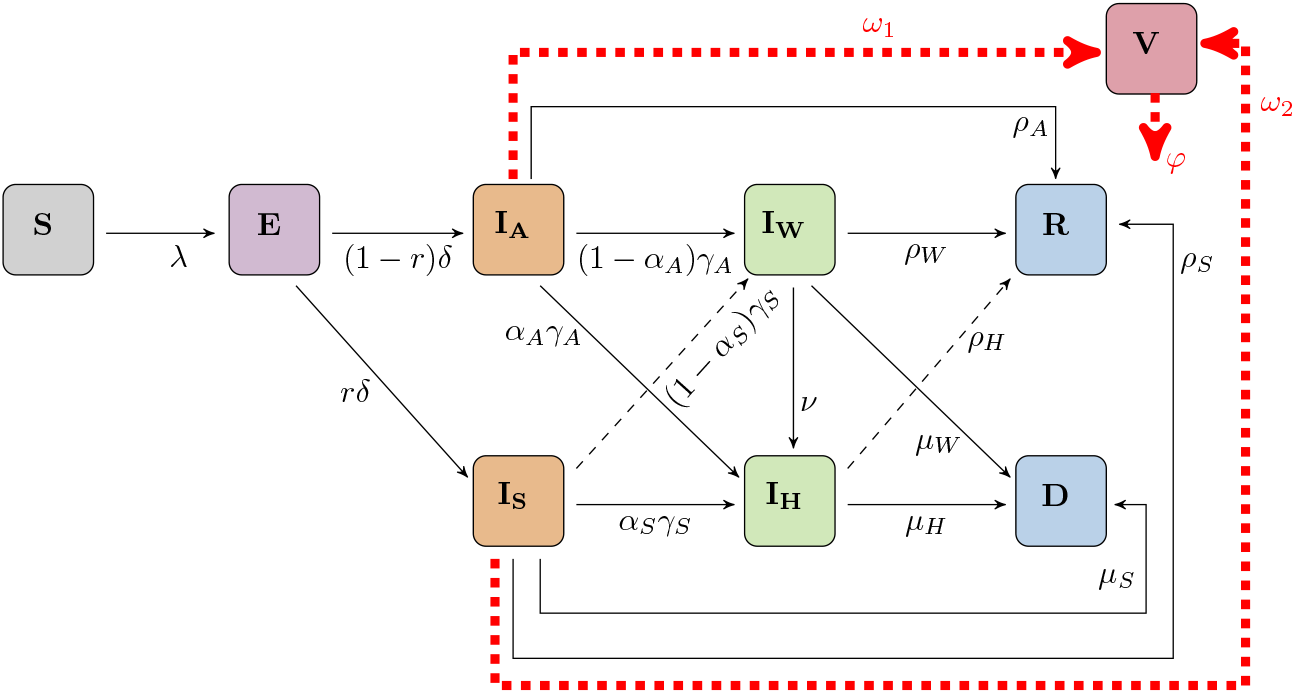
Diagram of the SEIRVD model with direct and indirect transmission of COVID-19.

The model diagram presented in Figure 1 is described by the system of non-linear differential equations in Equation 1. In this model, *β*_*DA*_ and *β*_*DS*_ are the direct transmission rates for asymptomatic and symptomatic individuals respectively. The rate *β*_*DA*_ is the product of the probability of the COVID-19 transmission from a contact between individuals in *S* and in *I*_*A*_, and the number of contacts per day per individual. And the rate *β*_*DS*_ denotes the product of the probability of the COVID-19 transmission from a contact between individuals in *S* and in *I*_*S*_, and the number of contacts per day per individual. The transmission rate *β*_1_ represents the indirect transmission rate, which is the probability of the disease transmission from a contact between individuals in *S* and the virus in the environment or contaminated surfaces *V*, and the number of contacts per day per length.

The parameter *p*_1_ represents the proportion of susceptible individuals protected from direct transmission as a result of effective mask usage, while *p*_2_ is the proportion of susceptible individuals protected from indirect transmission (i.e. through environmental protection, such as sanitation, cleaning of surfaces and door handles, environmental disinfection, fumigation and so on). Exposed individuals become infectious at a rate *δ*, where a proportion *r* of these individuals are symptomatic, and the remaining (1 − *r*) are asymptomatic. Asymptomatic individuals are tested at the rate *γ*_*A*_, where a fraction *α*_*A*_ become hospitalized, and the remaining 1 − *α*_*A*_ are isolated. We believe that asymptomatic individuals show mild or no symptoms, but a small proportion of those tested who are more vulnerable could be hospitalized as a precautionary measure. Hence our justification for assuming that a fraction *α*_*A*_ of this population could be hospitalized. Similarly, symptomatic individuals are tested at the rate *γ*_*S*_, where a fraction *α*_*S*_ become hospitalized, and the remaining 1 − *α*_*S*_ are isolated. Asymptomatic, symptomatic, hospitalized and isolated individuals recover from the disease at rates *ρ*_*A*_, *ρ*_*S*_, *ρ*_*H*_ and *ρ*_*W*_, respectively. In addition, symptomatic, hospitalized and isolated individuals die at rates *µ*_*S*_, *µ*_*H*_, and *µ*_*W*_, respectively, ignoring deaths of asymptomatic individuals since there are no available data.

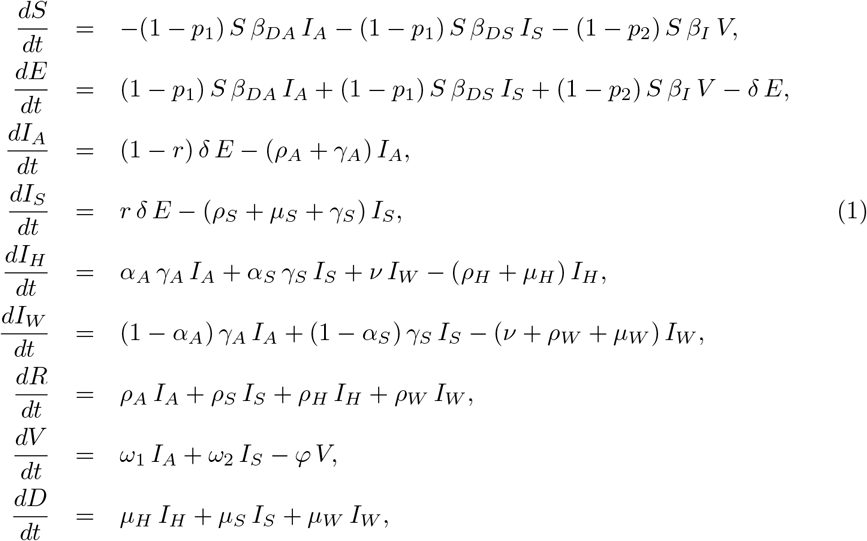

where *S*(0) = *S*_0_, *E*(0) = *E*_0_, *I*_*A*_(0) = *I*_*A*0_, *I*_*S*_(0) = *I*_*S*0_, *I*_*H*_ (0) = *I*_*H*0_, *I*_*W*_ (0) = *I*_*W* 0_, *R*(0) = *R*_0_ = 0, *D*(0) = *D*_0_ = 0, *V* (0) = *V*_0_, in a population of constant total size *N* (0) = *N*_0_, where *N* (0) as described in Table (1) is given as

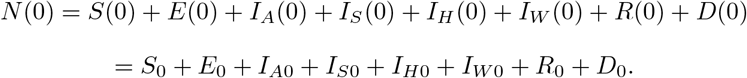

It is possible that some isolated individuals develop complications due to some pre-existing condition when exposed to the virus. Our model therefore assumes that isolated individuals with degenerating health condition are moved to the hospital at a rate *ν*. It is well-known that even though asymptomatic individuals show no symptoms, they still shed the virus and transmit infection. In our model, asymptomatic and symptomatic individuals shed viruses at rates *ω*_1_ and *ω*_2_, respectively. Viruses shed lead to new infections when susceptible individuals come in contact with them. The decay rate of the virus is *φ*.

### Risk assessment with or without indirect transmission

The following two subsections give the analytical computation of the effective reproduction number and the final size relation for the SEIRVD model (1) to assess the risk of COVID-19 epidemic with and without indirect transmission.

### Effective reproduction number

The basic reproduction number ℛ _0_ of a disease is defined as the number of secondary infections produced by one infectious individual introduced into a totally susceptible population. The reproduction number estimated after the introduction of intervention measures is called the *effective reproduction number*. In this model, the number of susceptible individuals that will be infected is reduced by *p*_1_ (proportion protected from direct transmission) and *p*_2_ (proportion protected from indirect transmission), and the reproduction number computed is called the effective reproduction number ℛ _*e*_. Using the next generation matrix approach [26, 27] as in [28–30], we have the effective reproduction number given as

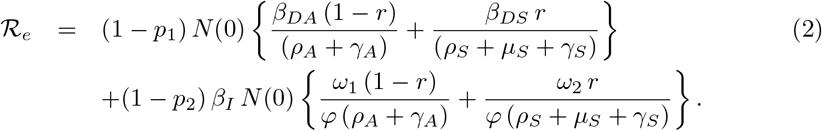

Note that *N* (0) = *N* since the total population is constant at all time. ℛ_*e*_ can also be written as ℛ_*e*_ = ℛ_*D*_ + ℛ_*I*_, where

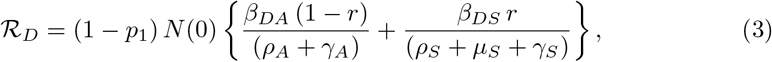

and

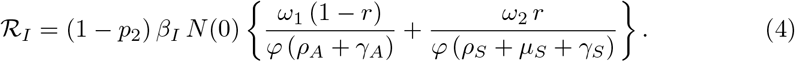

The expression for ℛ_*e*_ in equation (2) denotes the secondary infections contributed by direct transmission ℛ_*D*_ and indirect transmission ℛ_*I*_.

### The final size relation

The final epidemic size can be derived from the solution of the final size relation. This relation gives an estimate of the total number of infections and the epidemic size for the period of the epidemic using the parameters in the model [28, 31, 32]. Therefore, in order to estimate the total number of cases and deaths, we use the approach in [28–30] to derive the final size relation as

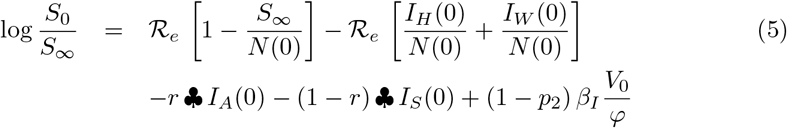

where

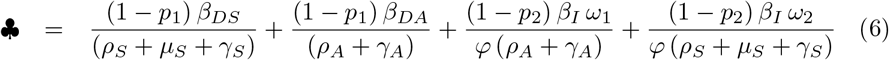

Equation (5) implies *S*_∞_ *>* 0.

Equation (5) is called the final size relation, and gives the relationship between the reproduction number ℛ _*e*_ and the final epidemic size. The total number of infected population over the course of the epidemic is given by *N* (0) − *S*_∞_, which can be described in terms of the attack rate as 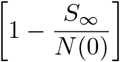.

### Important conclusions to inform the risk

We can epidemiologically interpret the reproduction number ℛ_*e*_ as the addition of the average number of new cases of COVID-19 generated by individuals in the classes *I*_*A*_ and *I*_*S*_ through direct contact (ℛ_*D*_) and indirect contact (ℛ_*I*_). In addition, we can interpret ℛ_*D*_ and ℛ_*I*_ as follow.

1. The reproduction number for direct transmission (ℛ_*D*_): the first term in the expression of equation 3 represents the product of the transmission rate of asymptomatic infectious individuals in the *I*_*A*_ class (*β*_*DA*_), the proportion of individuals becoming asymptomatic ((1 −*p*_1_)*N* (0)(1 − *r*)), and the average duration spent in the *I*_*A*_ class 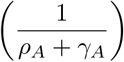, while the second term represents the product of the transmission rate of symptomatic infectious individuals in the *I*_*S*_ class (*β*_*DS*_), the proportion of individuals becom(ing symptomatic ((1 − *p*_1_)*N* (0)*r*), and the average duration spent in the *I*_*S*_ class 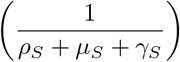
2. The reproduction number for indirect transmission (ℛ_*I*_): the first term in the expression of equation 4 represents secondary infections caused indirectly through the virus since a single infective *I*_*A*_ sheds a quantity *ω*_1_ of the virus per unit time for a time period 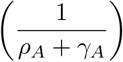 and this virus infects *β*_*I*_(1 − *p*_2_)*N* (0)(1 − *r*) susceptible individuals per unit time for a time period 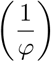, while the second term represents secondary infections caused indirectly through the virus since a single infective *I*_*S*_ sheds a quantity *ω*_2_ of the virus per unit time for a time period 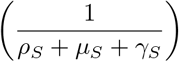and this virus infects *β*_*I*_ (1 − *p*_2_)*N* (0)*r* susceptible individuals per unit time for a time period 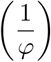.

The following easily proved theorem will give the summary of the benefit of the reproduction number *ℛ*_*e*_.

#### Theorem 1

*For system* (1), *the infection dies out whenever* ℛ_*e*_ *<* 1 *and epidemic occur whenever* ℛ_*e*_ *>* 1.

For the final epidemic size, we have that

1. If the outbreak begins through direct contact and some infected individuals (*V*_0_ = 0), the final size relation takes the form

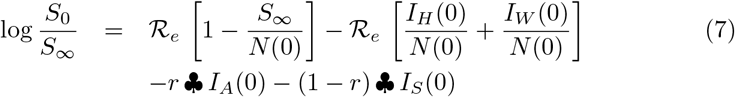
2. If the outbreak begins through direct contact and no infected individuals (*I*_*A*_ = *I*_*S*_ = *I*_*H*_ = *I*_*W*_ = 0, *V*_0_ = 0), the final size relation takes the form

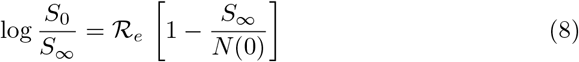
3. If the outbreak begins through direct and indirect contact and no infected individuals (*I*_*A*_ = *I*_*S*_ = *I*_*H*_ = *I*_*W*_ = 0), the final size relation takes the form

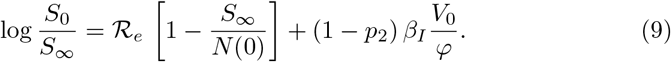

### Modeling scenarios

For model fitting, the initial values are set to the scenario in the data obtained from [33] (see scenario 1 on Table 6). Prior to March 1, the COVID-19 cases in Toronto were imported and not due to local transmission of the virus. Hence the data from March 1 onward was used for parameter estimation and model simulations. For model calibration, some of the parameters are adjusted to changes in policy as public health interventions. Hence, we subdivided some parameters (*β*_*DA*_, *β*_*DS*_, *β*_*I*_, *p*_1_, *p*_2_, *ν*) into stages I, II and III. Stage I is the time from March 1 to 12 (the time when the stay-at-home policy was implemented), stage II is the time from March 12 to May 19 (the time when reopening began) and stage III is the time from May 19 to the end of the data (August 31). We believe that change in policy will significantly influence behavioural changes and estimating these parameters for different stages will account for this. It is worth noting that no parameter for direct transmission was examined in this study since different studies have shown the effectiveness of NPIs for direct transmission models.

### Main Outcomes

We give numerical results of the SEIRVD model (1) computed using the MATLAB numerical ODE solver ODE45 [34]. These simulations and analytical show the cumulative number of cases and the prevalence of symptomatic individuals over time for three different scenarios. In addition, we present contour plots of the final epidemic size with respect to parameters of the model. We compare different scenarios of virus shedding on the peak, peak size, final epidemic size, the effective reproduction numberℛ_*e*_ by varying the two parameters *ω*_1_ and *ω*_2_ to estimate the impact of indirect transmission on COVID-19 epidemic in settings like Toronto. In addition, we estimated the cumulative cases and prevalence of symptomatic individuals for stages I, II, III to know the intensity of the effect of shedding at different stage of the epidemic. To estimate the level of shedding that could reduce indirect transmission and eventually decrease or eliminate the COVID-19 epidemic, we compare the outcomes with and without indirect transmission.

## Results

### Parameter estimation

Some of the parameters of the model (1) are assumed and derived from the literature (see Table 3), while others are estimated from the MCMC parameter estimation (see Table 2) using the cumulative confirmed COVID-19 cases by episode date in Toronto from March 1, 2020 to August 31, 2020 (Data from Toronto Public health)

**Table 1.**
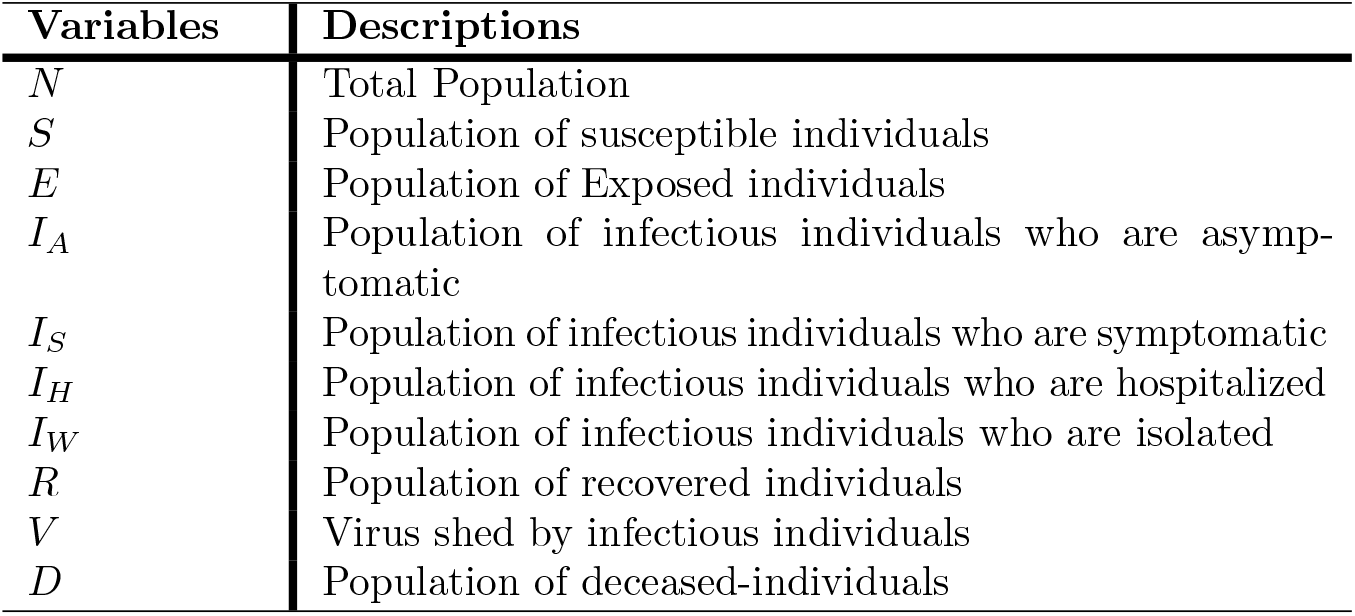
Model variables and descriptions.

| Variables | Descriptions |
| --- | --- |
| $N$ | Total Population |
| $S$ | Population of susceptible individuals |
| $E$ | Population of Exposed individuals |
| $I_A$ | Population of infectious individuals who are asymptomatic |
| $I_S$ | Population of infectious individuals who are symptomatic |
| $I_H$ | Population of infectious individuals who are hospitalized |
| $I_W$ | Population of infectious individuals who are isolated |
| $R$ | Population of recovered individuals |
| $V$ | Virus shed by infectious individuals |
| $D$ | Population of deceased-individuals |

**Table 2.** Estimated parameters of the model (1) in stages I, II, and III.

| Parameters | Descriptions | Estimated Mean Values<br>(Stages: I, II, III) | Sources |
| --- | --- | --- | --- |
| $\beta_{DS}$ | Transmission rate between $S$ and $I_S$ | I: 4.306e−07 person<sup>−1</sup> day<sup>−1</sup><br>II: 3.639e−07 person<sup>−1</sup> day<sup>−1</sup><br>III: 5.953e−07 person<sup>−1</sup> day<sup>−1</sup> | MCMC |
| $\beta_{DA}$ | Transmission rate between $S$ and $I_A$ | I: 6.220e−08 person<sup>−1</sup> day<sup>−1</sup><br>II: 1.598e−07 person<sup>−1</sup> day<sup>−1</sup><br>III: 5.778e−08 person<sup>−1</sup> day<sup>−1</sup> | MCMC |
| $\beta_I$ | Transmission rate for indirect (Environmental transmission) between $S$ and $V$ | I: 1.6462e−07 length<sup>−1</sup> day<sup>−1</sup><br>II: 1.0618e−07 length<sup>−1</sup> day<sup>−1</sup><br>III: 6.814e−09 length<sup>−1</sup> day<sup>−1</sup> | MCMC |
| $p_1$ | Proportion of susceptible individuals effectively using mask (masks usage) | I: 0.24701<br>II: 0.89787<br>III: 0.9865 | MCMC |
| $p_2$ | Proportion of individuals protected from environmental transmission | I: 0.30844<br>II: 0.77061<br>III: 0.55297 | MCMC |
| $\mu_S$ | Disease induced death rates for symptomatic individuals in the compartments $I_S$ | 0.034632 day<sup>−1</sup> | MCMC |
| $\rho_S$ | Recovery rate of symptomatic ($I_S$) individuals | 0.019475 day<sup>−1</sup> | MCMC |
| $\gamma_A$ | Testing rate of $I_A$ individuals | 0.029389 day<sup>−1</sup> | MCMC |
| $\gamma_S$ | Testing rate of $I_S$ individuals | 0.012072 day<sup>−1</sup> | MCMC |
| $\nu$ | Hospitalization rate of isolated individuals | I-II: 0.13819 day<sup>−1</sup><br>III: 0.012271 day<sup>−1</sup> | MCMC |
| $\alpha_A$ | Proportion of asymptomatic individuals who became hospitalized after testing | 0.74087 | MCMC |
| $\alpha_S$ | Proportion of symptomatic individuals who became hospitalized after testing | 0.82189 | MCMC |
| $\omega_2$ | Virus shedding rate for infectious individuals in the compartment $I_S$ (The rates of viral release into the environment by symptomatic individuals) | 0.025193 person<sup>−1</sup> day<sup>−1</sup> ml<sup>−1</sup> | MCMC |

**Table 3.**
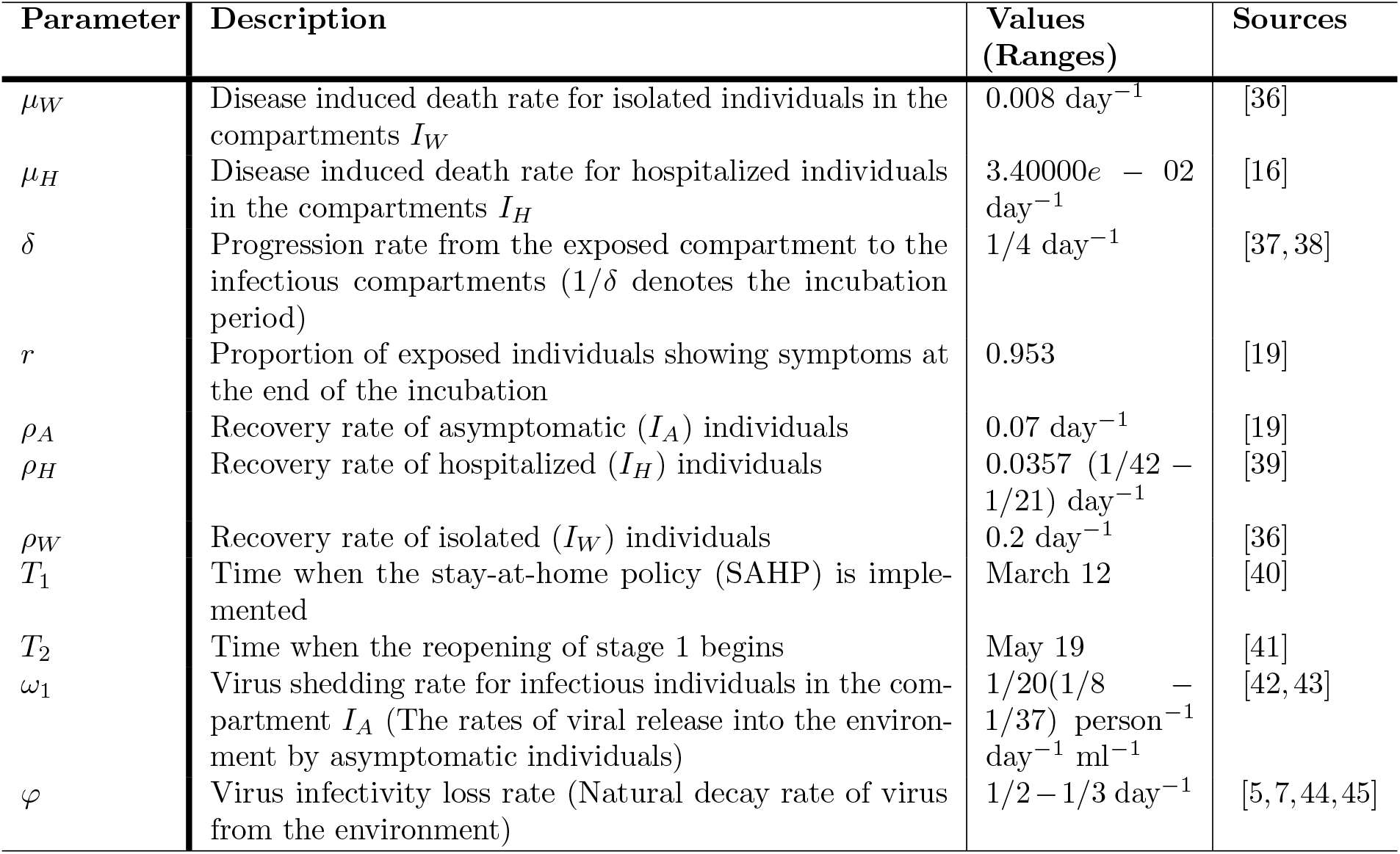
Model parameters.

We estimated 13 unknown parameters, namely the direct and indirect transmission rates (*β*_*DA*_, *β*_*DS*_, *β*_*I*_), proportion of effective mask usage (*p*_1_), proportion of individuals protected from indirect transmission (*p*_2_), death and recovery rate for symptomatic individuals (*µ*_*S*_, *ρ*_*S*_), testing rates for asymptomatic and symptomatic individuals (*γ*_*A*_, *γ*_*S*_), hospitalization rate of isolated individuals, asymptomatic and symptomatic individuals (*ν, α*_*A*_, *α*_*S*_), and shedding rate of symptomatic individuals (*ω*_2_), using Bayesian methods. For the parameter estimation, we used the multivariate Gaussian distribution as priors for the parameters, and the mean of the posterior distributions as the estimated parameters. We use Markov Chain Monte Carlo (MCMC) methods and employ the adaptive Matropolis-Hasting algorithm [35]. We assess the chain convergence by the Geweke statistic, with all values seen to be greater than 0.95, and the goodness of fit by the Normalized Mean Square Error (NMSE) of 0.98. The Geweke statistic indicates a satisfactory chain convergence, while the NMSE shows that the model fits the data very well as shown in Fig 2. The estimated parameters are shown in Table 2.

**Fig 2.**
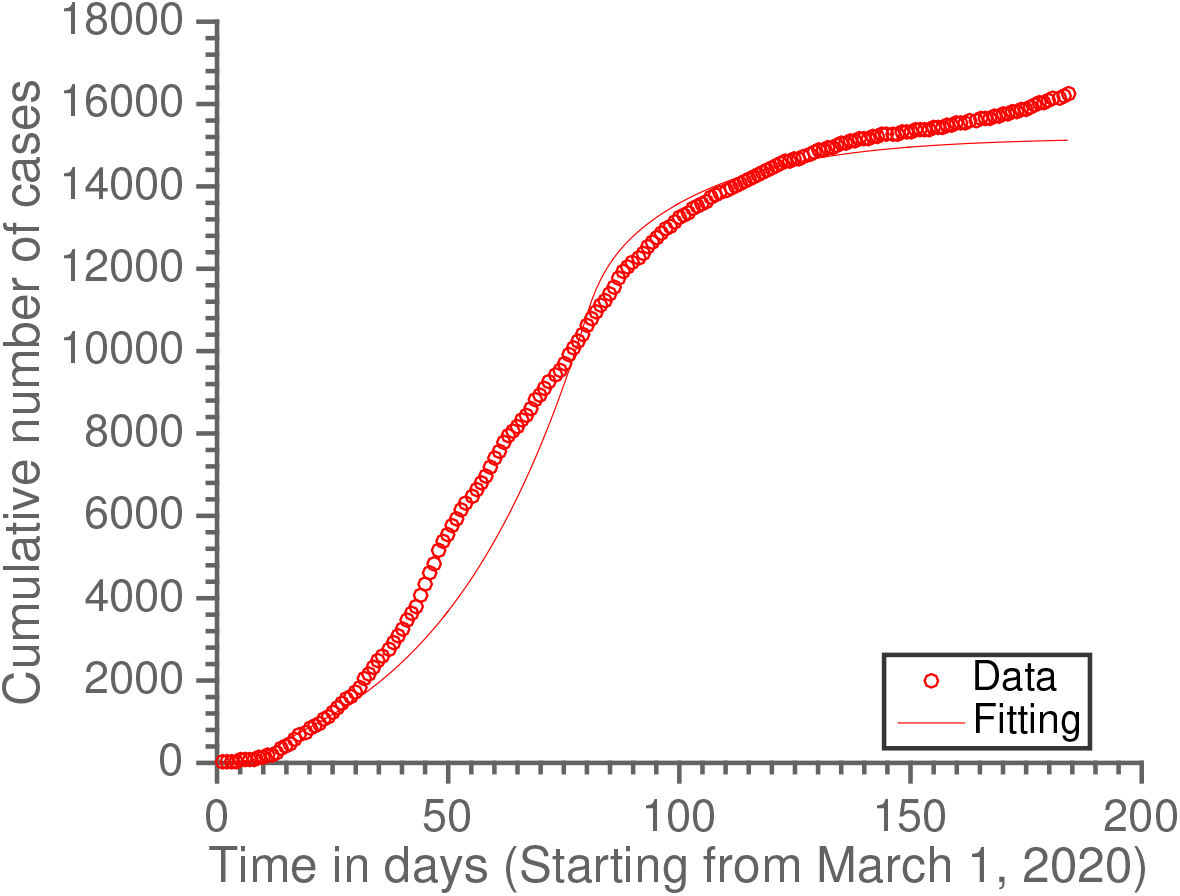
Toronto Data and estimate of the cumulative number of confirmed cases from March 1 to August 31, for model (1). Parameters used are given in Tables 2 and 3.

The results of the parameter estimation as in Table 2 indicate that most reported cases in Toronto from March to August are reportedly through direct transmission since the estimated shedding rate for symptomatic individuals is very low with *ω*_2_ = 0.025193 (see Table 2). This shows that based on our model outcome from March 1 to August 31, indirect transmission seem to have little effect on the number of confirmed cases in Toronto. Therefore, we explore scenarios for when *ω*_2_ will actually impact the epidemic (i.e. when *ω*_2_ ≥ 0.5) and show the potential effect of indirect transmission. Hence, in the following subsection, we explore different situations in our numerical simulations when *ω*_2_ = 0.5 or varied.

### The reproduction number for stages I, II and III using Toronto related scenario

In this section, we show the impact of the parameters estimated at different stages of the epidemic on the reproduction number for Toronto scenario. Table 4 and 5 show estimates of the reproduction number using three different stages for Toronto scenario. In Table 4 the estimated parameter for the symptomatic shedding rate (*ω*_2_ = 0.025193) is used, while *ω*_2_ = 0.5 is used in Table 5 for illustrative purpose. Other parameters used are as given in Tables 2 and 3.

**Table 4.** The reproduction number for stages I, II and III using Toronto related scenario with *ω*_2_ = 0.025193.

| Stages | Direct transmission<br>($\mathcal{R}_D$) | Indirect transmission<br>($\mathcal{R}_I$) | $\mathcal{R}_e$ ($\mathcal{R}_D + \mathcal{R}_I$) |
| --- | --- | --- | --- |
| Stage I | 13.87 | 0.26 | 14.13 |
| Stage II | 1.60 | 0.06 | 1.66 |
| Stage III | 0.34 | 0.01 | 0.35 |

**Table 5.** The reproduction number for stages I, II and III using Toronto related scenario with *ω*_2_ = 0.5.

| Stages | Direct transmission<br>($\mathcal{R}_D$) | Indirect transmission<br>($\mathcal{R}_I$) | $\mathcal{R}_e$ ($\mathcal{R}_D + \mathcal{R}_I$) |
| --- | --- | --- | --- |
| Stage I | 13.87 | 4.8620 | 18.73 |
| Stage II | 1.60 | 1.4 | 2.65 |
| Stage III | 0.34 | 0.13 | 0.47 |

**Table 6.** Results for different scenarios of initial condition used for model 1.

| Scenarios | Initial conditions (References) | Transmission routes | Number of symptomatic individuals at the peak | Peak time | Final size |
| --- | --- | --- | --- | --- | --- |
| **Scenario 1:** |  |  |  |  |  |
| Toronto related scenario | $N(0) = 2956024$ ([46]),<br>$S(0) = N(0) - 21$, $E(0) = 0$,<br>$I_A(0) = 0$, $I_S(0) = 11$ ([33]),<br>$I_H(0) = 0$, $I_W(0) = 0$, $R(0) = 10$ ([33]), $D(0) = 0$ ([33]),<br>$V(0) = 0$ | Direct & Indirect | $5.7109e + 04$ | 83 | $2.0725e + 05$ |
| | | Direct | $3.6528e + 04$ | 84 | |
| **Scenario 2:** |  |  |  |  |  |
| No initial infected population and $V_0 = 5$ | $N(0) = 2956024$, $S(0) = N(0)$, $E(0) = 0$, $I_A(0) = 0$, $I_S(0) = 0$, $I_H(0) = 0$,<br>$I_W(0) = 0$, $R(0) = 0$, $D(0) = 0$, $V(0) = 5$ | Direct & Indirect | $4.0681e + 03$ | 84 | $1.5143e + 04$ |
| | | Direct | $2.6064e + 03$ | 84 | |
| **Scenario 3:** |  |  |  |  |  |
| A few initial infected population and $V_0 = 5$ | $N(0) = 2956024$, $S(0) = N(0) - 21$, $E(0) = 0$, $I_A(0) = 0$, $I_S(0) = 11$, $I_H(0) = 0$,<br>$I_W(0) = 0$, $R(0) = 10$, $D(0) = 0$, $V(0) = 5$ | Direct & Indirect | $6.0685e + 04$ | 83 | $2.1985e + 05$ |
| | | Direct | $3.8807e + 04$ | 84 | |

Here, we compare Tables 4 and 5 for when *ω*_2_ = 0.025193 and *ω*_2_ = 0.5, respectively. We see from these tables that the impact of indirect transmission is lower in Toronto due to lower amount of viruses being shed by symptomatic individuals (*ω*_2_ = 0.025193). Using Table 5 for illustration, we have that the reproduction number changing and increasing when more viruses are being shed. For both tables, even though the contribution of ℛ_*D*_ to the total ℛ_*e*_ seems higher than ℛ_*I*_, the value of ℛ_*e*_ is still able to increase with increase in *ω*_2_. The full description of how changes in *ω*_2_ affects the reproduction number ℛ_*e*_ is shown in Figure 5.

**Fig 3.**
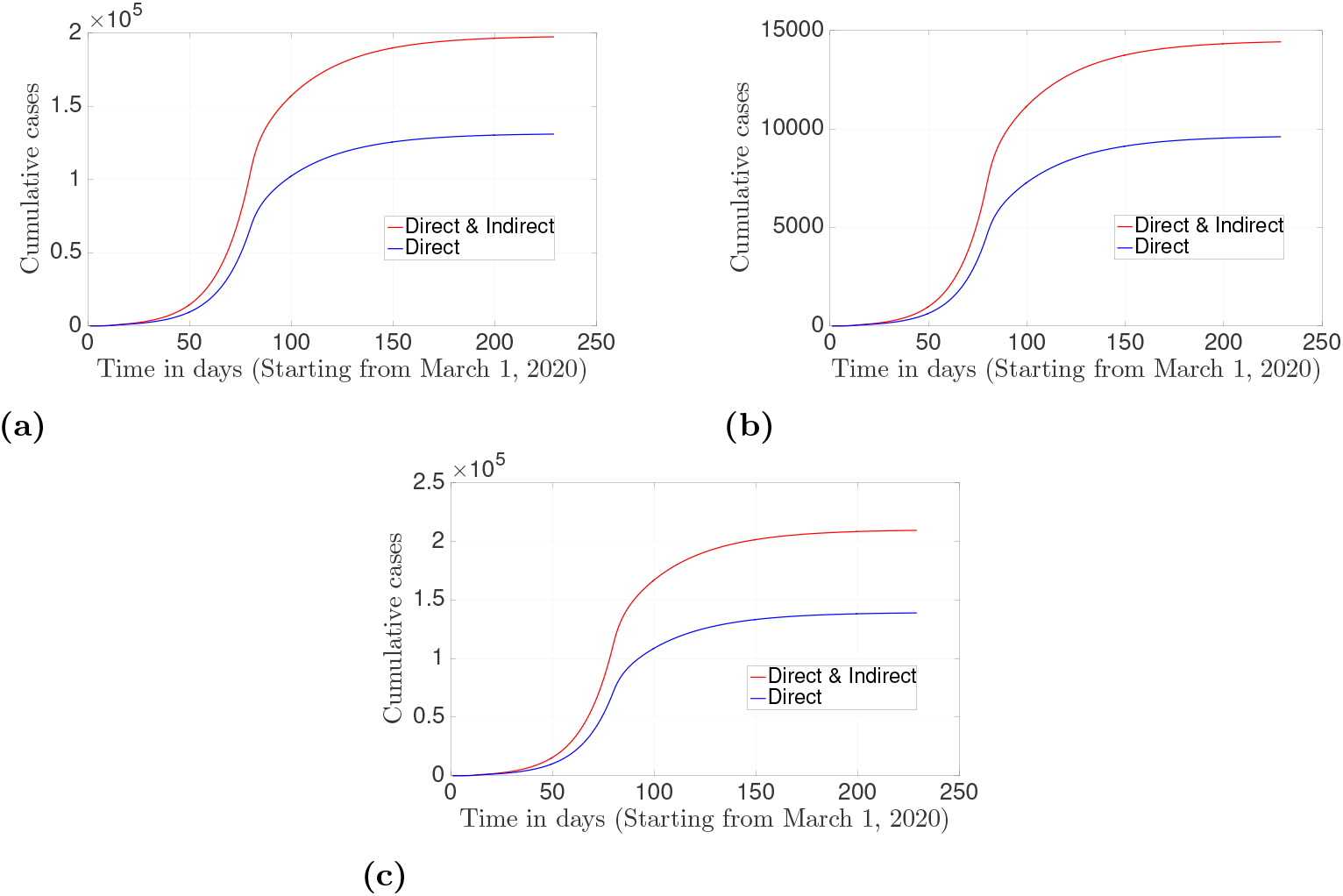
Cumulative confirmed cases over time for the model (1) for both direct and indirect transmission routes (red) and only direct transmission route (blue) using (a) Scenario 1, (b) Scenario 2, and (c) Scenario 3 in Table 6. The parameters used are given in Tables 2 and 3 except for ω_2_ = 0.5.

**Fig 4.**
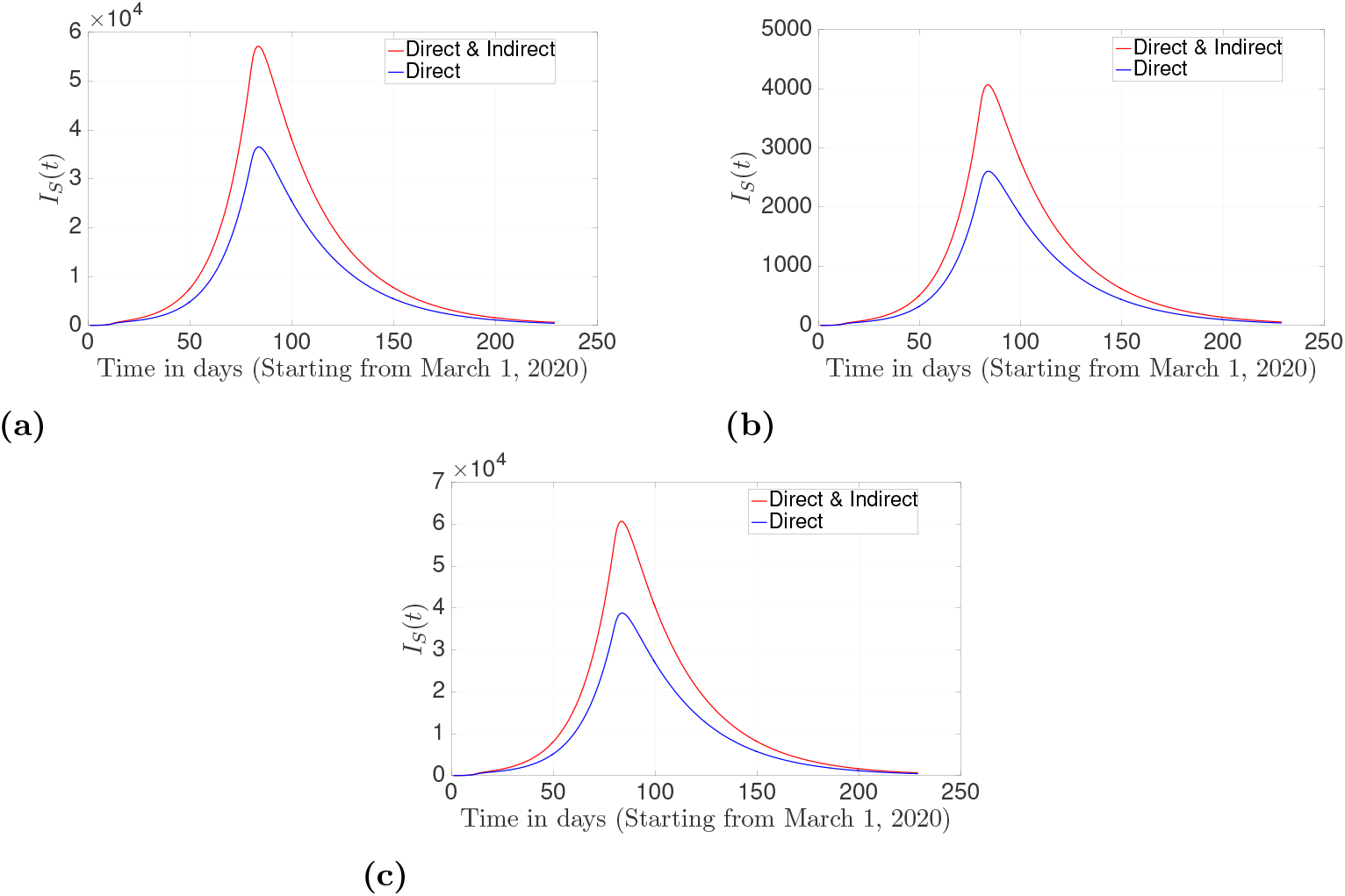
Prevalence of symptomatic individuals over time for the model (1) for both direct and indirect transmission routes (red), and only direct transmission route (blue) using (a) Scenario 1, (b) Scenario 2, and (c) Scenario 3 in Table 6. The parameters used are given in Tables 2 and 3 except for ω_2_ = 0.5.

**Fig 5.**
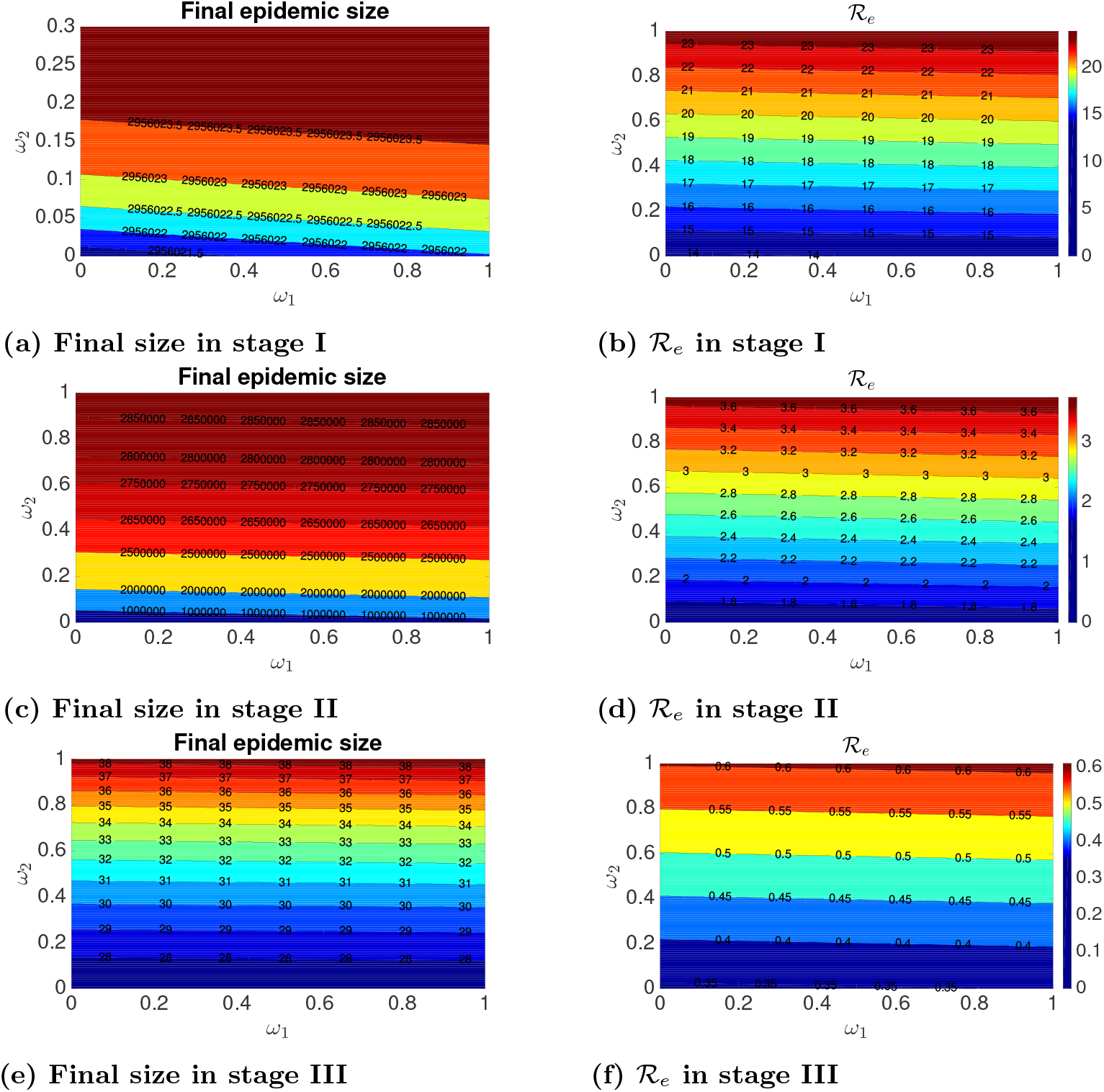
Contour plots of the final epidemic sizes (column 1) and the reproduction number _e_ (column 2) for the model (1) using parameters in stages I, II, and III from Table 2, for the Toronto related scenario. Column 1 which consists of Figs 5a, 5c, and 5e are showing the effect of varying the shedding rates ω_1_ and ω_2_ on the final epidemic sizes for stages I, II, and III respectively, while column 2 which consist of Figs 5b, 5d, and 5f are showing the effect of varying the shedding rates ω_1_ and ω_2_ on the reproduction number _e_ stages I, II, and III respectively. Other parameters are as given in Tables 2 and 3. Rows 1, 2 and 3 are for stages I, II and III, respectively.

For both direct and indirect transmission, the reproduction numbers are highest in stage I and lowest in stage III, which means that infection is highest in stage I. We can interpret *biologically* that if interventions and control measures in stage I are kept the same throughout the outbreak, the epidemic peak will be quickly reached within a shorter peak time and with a higher reproduction number compared to other stages.

### Cumulative confirmed cases and prevalence of symptomatic individuals

Table 6 shows three different scenarios with initial conditions, and different estimates of the number of symptomatic individuals.

Numerical simulations of the SEIRVD model (1) for the scenarios described in Table 6 are presented in Fig 3 and Fig 4. Fig 3 shows the cumulative number of cases over time, while Fig 4 shows the corresponding prevalence of symptomatic individuals for the three different scenarios considered. For the results in these figures, the red curves are for the cases where both direct and indirect transmission occurs (double transmission route), and the blue curves are for direct transmission only (single transmission route). Figs 3a and 4a are for the Toronto related scenario (*Scenario* 1 *in Table 6*), where there are only a few infected individuals at the beginning of the epidemic, and no virus has been shed. In this scenario, the outbreak begins with direct transmission, and at the peak, there are approximately 57109 number of symptomatic individuals for the model with both Direct and Indirect transmission peaking at day 83, and 36528 for that with direct transmission only peaking at day 84 (See Table 6 and Fig 4a). The final size for this scenario is given as 207250.

For the scenario with *V*_0_ = 5 and no infected individuals at the beginning (*Scenario* 2 *in Table 6*), denoting that the outbreak begins with indirect transmission, there are about 4068 symptomatic individuals for the model with both direct and indirect transmission, and 2606 for the model with only direct transmission at the peak and both peaking at day 84 (See Fig 4b). The cumulative number of confirmed cases and the prevalence of symptomatic individuals for this scenario are shown in Figs 3b and 4b, respectively. In addition, the final size for this scenario is estimated as 15143, which is lower than the final size in scenario 1.

The cumulative number of cases and prevalence of symptomatic individuals for the scenario with a few infected individuals at the beginning of the epidemic and *V*_0_ = 5 (*Scenario* 3 *in Table 6*) are shown in Figs 3c and 4c, respectively. This scenario is used to model an instance where the epidemic starts with both transmission routes. Here, there are about 60685 symptomatic individuals at the epidemic peak, which occurs at day 83. For the case with only direct transmission, the epidemic peaked at day 84 with 38807 symptomatic individuals. The final size for this scenario is estimated as 219850, which is the highest of all scenarios.

We observe from the results presented in this section that no matter where and how the outbreak is starting, the case of direct and indirect transmission lead to more infections (almost doubling) than that of only direct transmission. This is due to the impact of indirect transmission which are not accounted for in the case with only direct transmission. In addition, we notice that scenario 2 has the lowest peak size and the same peak time. This is because there are no infected individuals in the population to shed viruses at the beginning of the epidemic. Note that no matter where the epidemic is starting (direct or indirect route for different scenarios), the disease is still able to spread but with different peak. But all scenarios in Table 6 show that the epidemic is much worse for scenario 3, i.e. the situation where the outbreak begins through direct and indirect transmission routes. In addition, we observe from our model outcome that the epidemic will reach its peak a day earlier (day 83) for direct and indirect transmission if we have some infected individuals at the beginning of the outbreak as in Scenarios 1 and 3. There seems to be no significant difference in the peak times of scenario 2 for both transmission routes.

### Effect of varying the shedding rates *ω*_1_ and *ω*_2_ on the final epidemic size and the effective reproduction number *ℛ*_*e*_

Here, we present contour plots of the final epidemic size and the effective reproduction number with respect to *ω*_1_ and *ω*_2_. The left panel of Fig 5 shows the final epidemic size of our model plotted with respect to the shedding rates *ω*_1_ and *ω*_2_ (Figs 5a, 5c and 5e in column 1). These figures are for parameters, interventions and control measures in stages I, II, and III, respectively as given in Table 2. We observe from these plots that the shedding rate of asymptomatic individuals (*ω*_1_) has no obvious effect on the final epidemic size for the three stages. On the other hand, as one may expect, an increase in the shedding rate of symptomatic individuals (*ω*_2_) increases the final epidemic size for all stages, especially for stage II since the data accounts for symptomatic individuals. The epidemic size is largest in stage I, reduced in stage II, and smallest in stage III. These results are shown in the first column of Figure 5.

The right panel of Fig 5 shows the contour plots of the reproduction number ℛ_*e*_ with respect to the shedding rates *ω*_1_ and *ω*_2_ (Figs 5b, 5d, and 5f in column 2) for stages I, II, and III described in Table 2. For all stages, we observe from these plots that the reproduction number increases as *ω*_2_ increases, and an increase in *ω*_1_ has no obvious effect, which is similar to the trend shown from the contour plots of the final epidemic sizes (column 1). This shows the impact of shedding of viruses in increasing cases since an increase in the value of *ω*_2_ means that more viruses are shed by symptomatic individuals, which denote more infections are caused through indirect transmission, thereby increasing the epidemic size as observed from the contour plots. In addition, we observe that stage III (Figs 5e and 5f) has the lowest final epidemic size and the reproduction number, denoting that if the parameters in stage III are kept during an outbreak, the disease is still able to spread but with a lower final size and reproduction number when compared to an outbreak with parameters in stages I and II. This *biologically* means that the shedding of the virus by symptomatic individuals has a greater impact on both final epidemic size and the reproduction number, and many models ignoring indirect transmission and viruses shed by this group will not account for this effect and may underestimate the true size of the epidemic. Furthermore, it also means that if different control measures and non-pharmaceutical interventions (NPIs) including lockdown, social distancing, mask usage were kept as in stage III, in addition to the effort to decrease indirect transmission by reducing the shedding rates, the disease will quickly die out (as seen from the reproduction number in stage III with low *ω*_1_ and *ω*_2_) and an increase in the final epidemic size will not be significant.

### Effect of varying the indirect transmission rate *β*_*I*_ and the proportion protected from environmental transmission *p*_2_ on the final epidemic size and the reproduction number ℛ_0_ for stages I, II, and III

In this section, we explore the effect of varying the indirect transmission rate *β*_*I*_ and the proportion of individuals protected from indirect transmission *p*_2_, on the final epidemic size for stages I, II and III. In Fig 6, we present contour plots of the final epidemic size with respect to the parameters *β*_*I*_ and *p*_2_. Figs 6a, 6b, and 6c are for stages I, II and III, respectively as given in Table 2.

**Fig 6.**
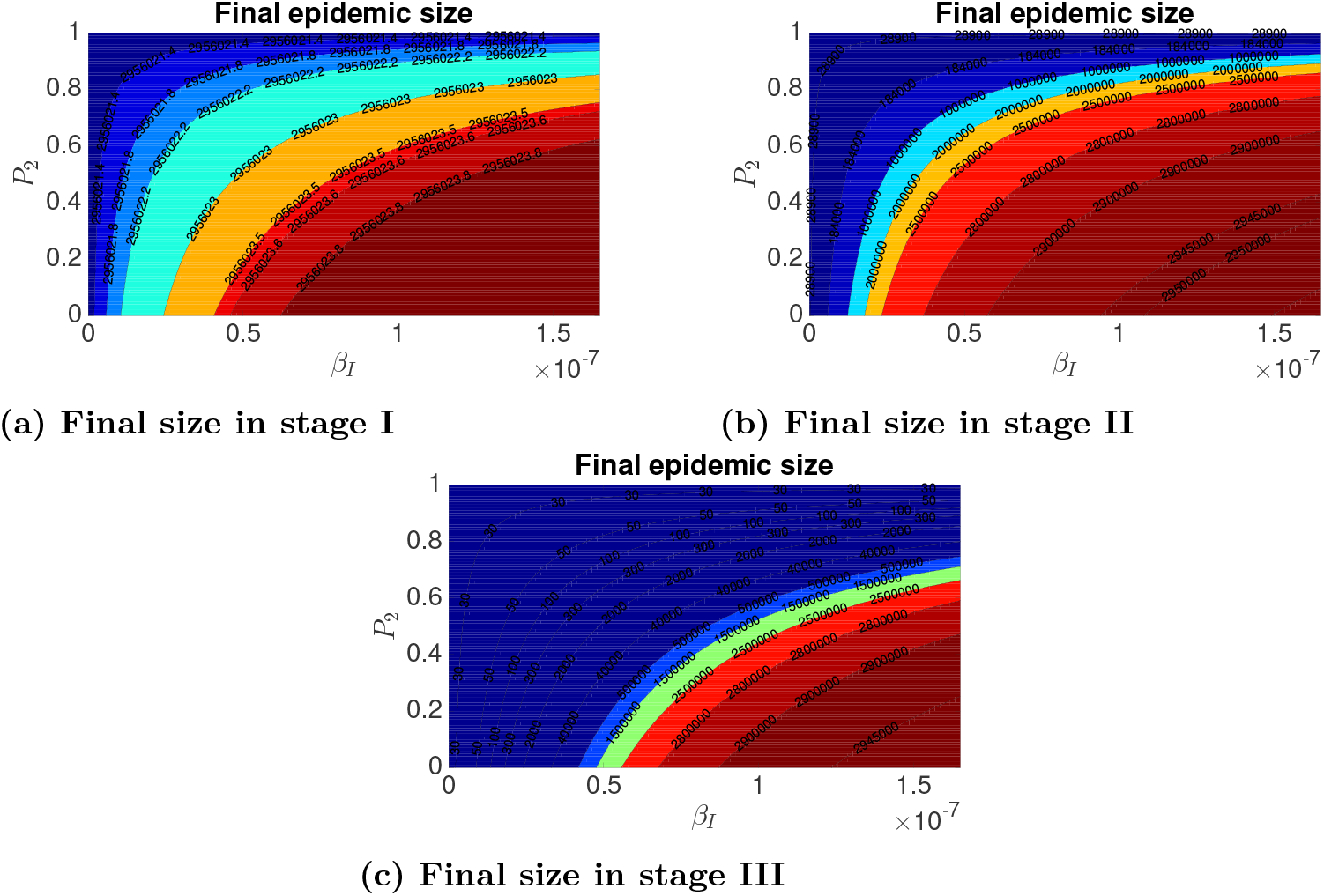
Contour plots of the final epidemic sizes for the model (1) for stages I, II, and III as specified in Table 2. Figs 6a, 6b, and 6c are for varying the indirect transmission rate β_I_ and the proportion of individuals protected from indirect transmission p_2_, with ω_2_ = 0.5 in stages I, II, and III, respectively. Other parameters are as given in Tables 2 and 3.

For all stages, we observe from these plots that the final epidemic size decreases as *p*_2_ increases, and increases as *β*_1_ increases. This shows the role of environmental protection in averting cases since an increase in the values of *p*_2_ means that more people are protected from contracting infections through indirect transmission, thereby lowering the epidemic size as observed from the contour plots. On the other hand, an increase in the values of *β*_*I*_ implies that infections are transmitted through indirect route at a higher rate, leading to a larger outbreak. In addition, we observe that stage III (Fig 6c) has lowest final epidemic sizes, followed by stage II, while stage I has highest final sizes. This means that if parameters and measures in stage III are implemented during an outbreak, the disease is still able to spread but with a lower final size when compared to other stages. We can *biologically* interpret that different control measures, especially measures related to indirect transmission (like *β*_*I*_ and *p*_2_) need to be implemented in order to curtail or eliminate the epidemic.

### Effect of varying the asymptomatic shedding rate *ω*_1_ on the prevalence of symptomatic individuals in stages I, II, and III

Next, we show the time dynamics of the prevalence of symptomatic individuals for different values of the asymptomatic shedding rate *ω*_1_, and for the three stages I, II, III in Table 2, for Toronto related scenario. Figs 7a, 7b and 7c are the prevalence of symptomatic individuals in stages I, II, and III, respectively. These results are generated for asymptomatic shedding rates *ω*_1_ = 0.01, 0.1, 0.1 and 1, with *ω*_2_ = 0.5. The remaining parameters are as given in Tables 2 and 3. The solid curves denote direct and indirect transmission routes (DI), while the dashed curves denote only direct transmission route (D). We observe from the results in stage I (Figure 7a) that the prevalence of symptomatic individuals do not change with increase in *ω*_1_ for the case of DI and D. Although, the peak in DI is always higher than that of D regardless of the value of *ω*_1_. This is because even though an increase in *ω*_1_ leads to more viruses being shed, the infection in stage I is already very high and therefore increasing the viruses being shed by asymptomatic individuals has no obvious effect on the prevalence of symptomatic individuals. Stage I has the highest number of symptomatic individuals at the peak, with a shorter peak time compared to stage II.

**Fig 7.**
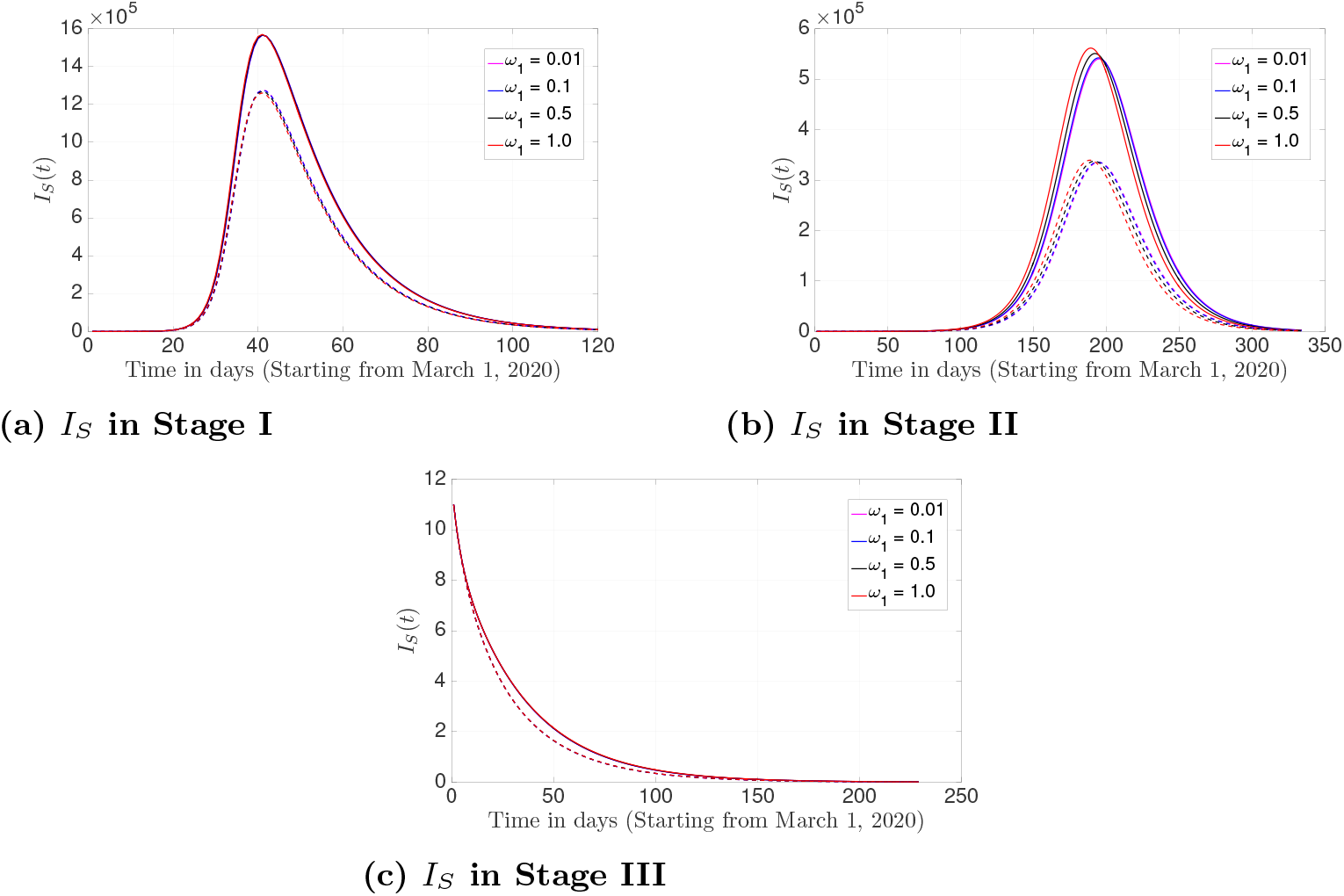
The prevalence of symptomatic individuals over time for the model (1) for both direct and indirect transmission routes (DI:solid curves) and only direct transmission route (D:dashed curves), generated for different values of the asymptomatic shedding rate ω_1_, with ω_2_ = 0.5 in stages I, II, and III. Other parameters are as given in Tables 2 and 3. Figs 7a, 7b and 7c are for stages I, II, and III respectively.

We also notice in stage II (Fig 7b) that the prevalence of symptomatic individuals at the peak increases slightly as *ω*_1_ increases for both DI and D with a lower number of symptomatic individuals at the peak, and with a longer peak time when compared to stage I. When parameters and measures in stage II are kept, the epidemic is delayed compared to stage I. In contrast to stages I and II, stage III (Fig 7c) shows that the prevalence of infected individuals decreases with time and the disease dies out. Also, we see no significant different in *I*_*S*_ for doth DI and D when *ω*_1_ is varied. This *biologically* means that the shedding of the virus by asymptomatic individuals has a slight impact on the transmission of infection for all stages, and many models ignoring transmission resulting from the viruses being shed may not account for this slight impact.

### Effect of varying the symptomatic shedding rate *ω*_2_ on the prevalence of symptomatic individuals in stages I, II, and III

Here we vary the shedding rate of symptomatic individuals *ω*_2_. Fig. 8 shows the prevalence of symptomatic individuals over time for stages I, II, and III in Table 2 for Toronto related scenario. The values of *ω*_2_ used are *w*_2_ = 0.1, 0.5, 0.7, 1.0, and the remaining parameters are as given in Tables 2 and 3. Figs 8a, 8b and 8c are for stages I, II, and III, respectively. Similar to our results and observations in Fig 7, from stage I (Figure 8a) and DI (solid curves), we observe that the prevalence of symptomatic individuals increases slightly as the shedding rate of symptomatic individual (*ω*_2_) increases. As *ω*_2_ increases, epidemic occurs faster, and quickly reaches it peak with an increasing number of infected individuals at the peak, and a decreasing peak time. In contrast, for stage I (Fig 8a) and D (Dashed curves), we observe that the prevalence of symptomatic individuals decreases slightly with an increase in *ω*_2_. As *ω*_2_ increases, both the number of symptomatic individuals at the peak and the peak time decrease. In order words, if parameters in stage I are implemented and more viruses are being shed by symptomatic individuals, we have more infections for DI and less infections for D. In addition, the difference in the peak of DI and D becomes wider as *ω*_2_ increases, showing how *ω*_2_ affects DI more compared to D. Stage I also buttresses our points and arguments from 7a that the peak in DI is higher than that of D regardless of the value of *ω*_2_, with DI accounting for the contribution of indirect transmission. Despite the differences in the peak and the peak time of DI and D in stage I, the epidemic using DI and D ended around the same time (at about 120 days)

**Fig 8.**
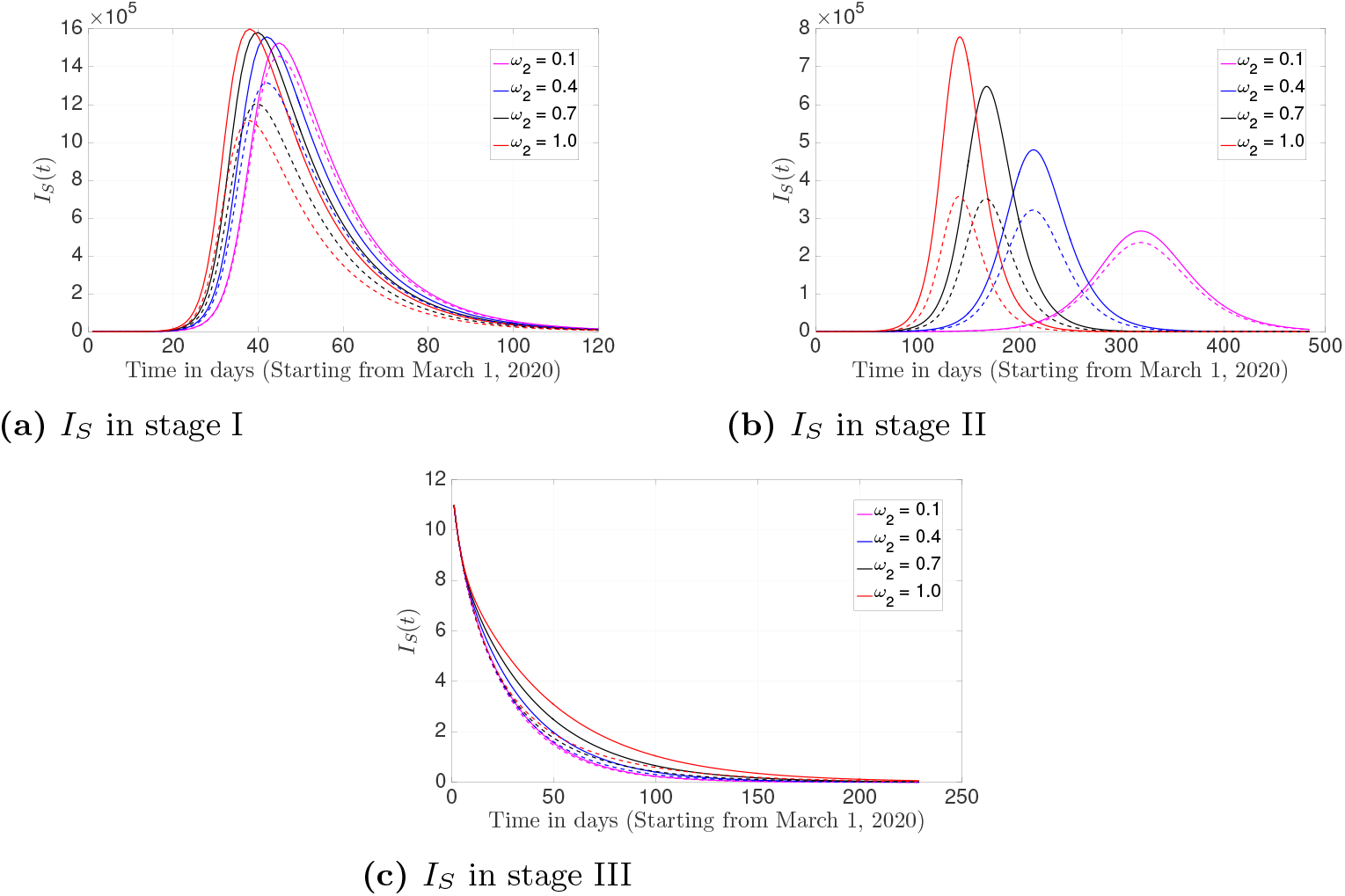
The prevalence of symptomatic individuals over time for the model (1) for both direct and indirect transmission routes (DI:solid curves) and only direct transmission route (D:dashed curves), generated for different values of the symptomatic shedding rate ω_2_ in stages I, II, and III. Other parameters are as given in Tables 2 and 3. Figures 8a, 8b and 8c are for stages I, II, and III respectively

In stage II (Fig 8b) and DI (solid curves), we observe that an increase in *ω*_2_ significantly increases the number of symptomatic individuals at the peak and with a decreasing peak time. Simply put, when *ω*_2_ increases, the infected individuals at the peak increases, the peak time decreases, meaning more infections which cause the epidemic to reach its peak at a shorter time. Similar trend is shown in stage II (Fig 8b) and D (dashed curves), except that an increase in *ω*_2_ slightly increases the number of symptomatic individuals at the peak. In addition, epidemic is delayed in stage II compared to stage I, and the effect of *ω*_2_ in stage II is significantly higher than that of stage I. Similar to our results in Fig 7c, *ω*_2_ = 1.0 accounted for more infections for DI, but the disease in this stage dies out within a shorter period of time. In For all values of *ω*_2_, and for DI and D in stage II, the epidemic started at about 100 days or latter, and ended at different time. But an increase in *ω*_2_ decreases the epidemic ending time (i.e., the lowest *ω*_2_ has the longest ending time). If *ω*_2_ is kept lower (for example, *ω*_2_ ≤ 0.1), the epidemic will occur late, but with a very low peak.

If parameters in stage III with different control measures are implemented the COVID-19 disease will die out within the shortest period of time compared to stages I and II. When these measures are implemented, the difference between DI and D may not be obvious. It is good to also note that varying *ω*_2_ has a greater effect on the peak size and peak time compared to varying *ω*_1_. This *biologically* means that the shedding of the virus by symptomatic individuals has a huge impact on both direct and indirect transmission (DI), especially when *ω*_2_ ≥ 0.4. Models with no indirect transmission and estimating the prevalence of symptomatic individuals, will account for the plots shown with dashed curves (D) without accounting for the differences in the solid curves (DI) and dashed curves (D). In other words, changes in *ω*_2_ impact the dynamics, the peak and the peak time more than changes in *ω*_1_.

### Effect of varying the symptomatic shedding rate *ω*_2_ on the peak size and peak time in stage II

Fig. 9 shows the peak size and peak time of symptomatic individuals in stages II. The values of *ω*_2_ used are *w*_2_ = 0.1, 0.5, 0.7, 1.0, and the remaining parameters are as given in Tables 2 and 3. Figs 9a and 9b denote the peak size and peak time respectively.

**Fig 9.**
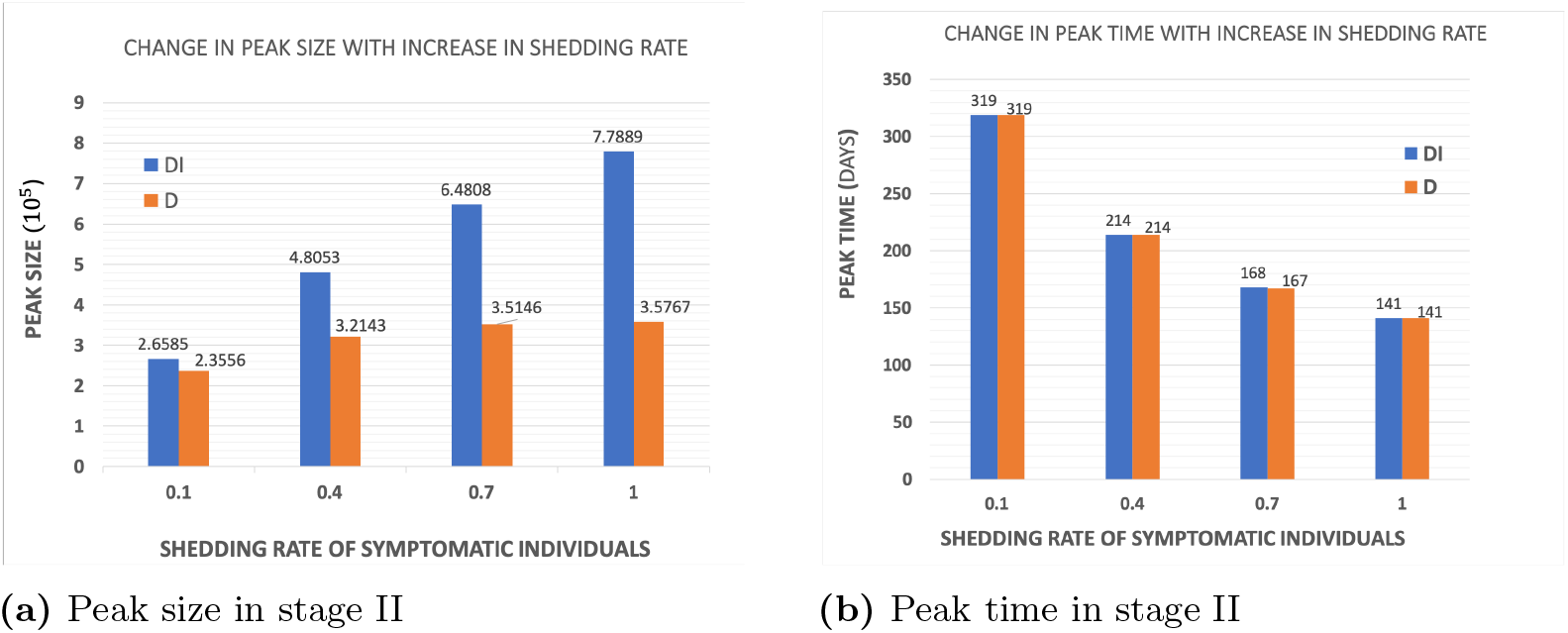
The peak size and peak time for the model (1) for both direct and indirect transmission routes (DI:blue) and only direct transmission route (D:red), generated for different values of the symptomatic shedding rate ω_2_ in stage II. Other parameters are as given in Tables 2 and 3. Figures 9a and 9b are for stage II

Similar to our results and observations in stage II of Fig 8, we observe that the peak size of symptomatic individuals increases as the shedding rate of symptomatic individual (*ω*_2_) increases for both DI and D. Simply put, when *ω*_2_ increases, the infected individuals at the peak size increases, the peak time decreases, meaning more infections which cause the epidemic to reach its peak at a shorter time. Although, the increase in peak size with increase in shedding rate is higher and more obvious in DI (blue bar) than in D (red bar) due to the impact of indirect transmission.

As *ω*_2_ increases, the peak time decreases and there seems to be no significant difference in the peak time of DI and D. In order words, if parameters and control measures in stage II are implemented and more viruses are shed by symptomatic individuals, we have increase in infections for both DI and D, but more for DI. In addition, the difference in the peak size of DI and D becomes wider as *ω*_2_ increases, showing how *ω*_2_ affects DI more compared to D. Furthermore, we see from 9a that the peak size in DI is higher than that of D regardless of the value of *ω*_2_, with DI accounting for the contribution of indirect transmission. In Fig. 9b, for all values of *ω*_2_, and for DI and D, the lowest *ω*_2_ has the longest peak time). If *ω*_2_ is kept lower (for example, *ω*_2_ ≤ 0.1), the epidemic will occur late and with a very low peak size.

From these simulations, it is obvious that difference and changes in control measures related to indirect transmission could cause changes in the peak size of the epidemic. In fact, they also have significant effect on the time of the peak and the end of the epidemic. This concludes the importance and effort needed to lower transmission occurring through indirect pathway by incorporating additional control and preventive measures.

## Discussion

We develop and analyze an ODE-based SEIRVD-type compartmental model. Several mathematical models have been developed to study and predict the transmission dynamics of the virus both locally within a specific region/country, and between multiple regions/countries [47, 48] without explicitly modeling the role of indirect transmission of SARS-CoV-2 on the epidemic, especially assessing how shedding of viruses impact the peak, peak time, effective reproduction number and the final epidemic size. Our study shows that the shedding of viruses, especially by symptomatic individuals has a huge effect on COVID-19 transmission, and efforts towards reducing virus shedding will significantly reduce the epidemic. We show that the epidemic of COVID-19 could be better curtailed when additional interventions relating to indirect transmission are combined with the current control measures. We see that reducing the shedding of SAR-CoV-2 to the minimum possible will reduce the peak, peak time, effective reproduction number and the final epidemic size.

Our results show larger epidemic peak and final sizes for the case of both direct and indirect transmission (DI) compared to that of only direct transmission (D), irrespective of the initial condition and the stage of the infection. The most successful strategy that give the lowest peak and the highest peak time for symptomatic individuals is to reduce the shedding rates of both asymptomatic and symptomatic individuals to lower than and 0.1 (i.e. *ω*_1_ *<* 0.01 and *ω*_2_ *<* 0.1) respectively. In addition, we show that the final epidemic size is highest when transmission rate (*β*_*I*_) is high and the rate of protection of individuals from environmental transmission (*p*_2_) is low. The final size decreases as indirect transmission rate decreases and protection increases, showing how implementing additional control measures could further reduce the epidemic. In general, we show that the elimination of COVID-19 may never be achieved if more viruses are shed and there are no interventions towards controlling the spread of infection through indirect transmission. Again, our results reinforce the fact that indirect transmission increases the epidemic of COVID-19. In other words, to eliminate the COVID-19 epidemic, indirect transmission cannot be ignored or neglected.

According to our model calibration, the shedding rate for symptomatic individuals from March to August in Toronto is very low. But for illustrative purposes, we used *ω*_2_ = 0.5 and varied *ω*_2_ where necessary to assess the potential impact of increasing shedding rates. For all level of shedding, we observe highest cases with DI when when compared to D, showing possible effects on indirect transmission. Our results seem consistent with existing studies on transmission of SARS-CoV-2 through aerosol and airborne [3–7, 10, 11].

As a limitation, the model is developed for a region or geographical area where the testing of individuals is readily available. This model does not consider heterogeneous mixing between two or more populations/regions. An extension of the model to include heterogeneous mixing of individuals consisting of multiple populations should be straightforward. Our work considered a general population and our results are generated as crude estimates. Projects on model stratification according to age, gender, region, socio-economic status are underway. We did not look into the impact of opinion and individual behaviour known to significantly modify the risk of COVID-19 transmission [49–51]. Therefore we could not evaluate the effect of indirect transmission for different opinion and behaviour, and the complexities that exist in behavioural changes. Even though, these conditions are somewhat important factors in modeling the transmission of COVID-19, modeling their effects would make the model more complicated, which is beyond the scope of this study, and therefore considered as one of the future works we intend to explore. Despite these limitations, our findings show that reducing the transmission of the virus through indirect route would significantly decrease the spread of the disease, especially when efforts are concentrated more on the symptomatic individuals through further isolation and/or hospitalization. We believe that models that do not consider indirect transmission of the virus may underestimate the cases, and consequently the final epidemic size and the reproduction number. In addition, models considering only direct transmission will overestimate the peak time since the peak time is reduced when more viruses are shed. Efforts such as fumigation, cleaning of surfaces, sanitation, towards reducing transmission from this route will help at lowering the epidemic of COVID-19. In addition, Toronto and other related cities need to introduce new disinfectant product to reduce indirect transmission.

## Conclusion

The use of SEIRVD-type model enables us to assess the impact of indirect transmission on COVID-19 epidemic. Our model is applicable to other similar settings to Toronto and would be great at measuring virus shedding like those considered in this study could impact COVID-19 transmission, dynamics and epidemic. Our work suggests the benefit and importance of some additional public health policies to address effect of shedding of viruses on COVID-19 epidemic, in Toronto and in other parts of the world. Given the current increase in reported cases, healthcare providers should ensure to further implement additional preventive and control measures to improve protection by increasing the physical distancing, fumigation, cleaning of surfaces, sanitation, disinfection, continue to promote mask usage, improve COVID-19 testing, and initiate immediate isolation of positive cases and their contacts when necessary. Successful implementation of these proceedings is crucial to addressing the COVID-19 epidemic.

## Data Availability

The data underlying the results presented in the study are available from Toronto Public Health

https://www.toronto.ca/home/covid-19/covid-19-latest-city-of-torontonews/covid-19-status-of-cases-in-toronto/

## Acknowledgments

This research was supported by the Canadian Institutes of Health Research (CIHR), Canadian COVID-19 Math Modelling Task Force, the Natural Sciences and Engineering Research Council of Canada (NSERC) and York University Research Chair program. The authors sincerely appreciate Toronto Public Health (TPH) for sharing their data on COVID-19 epidemic. Special thanks to Laboratory of Mathematical Parallel Systems (LAMPS) and Centre for Diseases Modeling (CDM) groups for discussion during model construction.

